## Supplementary material for "Child Anemia in Cambodia: A Descriptive Analysis of Temporal and Geospatial Trends and Logistic Regression-Based Examination of Factors Associated with Anemia in Children": S1 Table. Results of checking multicollinearity using Variance Inflation Factor

| **Variable** | **VIF** |
| --- | --- |
| Wealth quintile | 2.13 |
| Type of toilet facility | 1.96 |
| Children underweight | 1.63 |
| Birth order | 1.56 |
| Mother age | 1.50 |
| Children stunted | 1.45 |
| Mother education | 1.36 |
| Region | 1.24 |
| Children wasted | 1.2 |
| Place of residence | 1.18 |
| Recent diarrhea | 1.11 |
| Child age | 1.10 |
| Recent fever | 1.09 |
| Source of drinking water | 1.09 |
| Maternal anemia | 1.02 |
| Sex of child | 1.00 |
