## Supplementary material for "Child Anemia in Cambodia: A Descriptive Analysis of Temporal and Geospatial Trends and Logistic Regression-Based Examination of Factors Associated with Anemia in Children": S2 Table. Prevalence of Anemia in the 19 domains in CDHS 2005 2010 and 2014

| **Provinces** | **CDHS 2005 (n=2,933)** | | **CDHS 2010 (n=3,407)** | | **2014 (n=3,955)** | |
| --- | --- | --- | --- | --- | --- | --- |
|  | **%Anemia** | **95% CI** | **%Anemia** | **95% CI** | **%Anemia** | **95% CI** |
| Banteay Meanchey | 70.6 | (61.0-78.7] | 52.7 | (42.9-62.2] | 41.5 | (35.6-47.6] |
| Kampong Cham | 57.8 | (50.1-65.2] | 56.8 | (47.7-65.5] | 63.9 | (57.7-69.7] |
| Kampong Chhnang | 58.6 | (49.1-67.5] | 62.5 | (56.9-67.7] | 57.5 | (50.6-64.1] |
| Kampong Speu | 63.0 | (54.6-70.7] | 55.1 | (46.2-63.6] | 64.1 | (55.3-72.0] |
| Kampong Thom | 74.1 | (68.7-78.9] | 67.0 | (57.6-75.2] | 67.1 | (58.5-74.7] |
| Kandal | 57.2 | (48.4-65.5] | 56.5 | (46.6-65.9] | 59.8 | (50.1-68.8] |
| Kratie | 59.4 | (51.8-66.5] | 62.2 | (54.8-69.0] | 50.7 | (43.8-57.6] |
| Phnom Penh | 54.3 | (38.9-68.9] | 48.2 | (41.7-54.7] | 41.3 | (34.6-48.4] |
| Prey Veng | 57.0 | (48.1-65.5] | 51.6 | (43.3-59.9] | 54.5 | (44.7-64.0] |
| Pursat | **84.3** | (77.3-89.4] | 39.7 | (33.1-46.8] | 64.2 | (56.0-71.6] |
| Siem Reap | 78.0 | (70.5-84.0] | 61.4 | (54.3-68.0] | 50.5 | (44.0-56.9] |
| Svay Rieng | 68.0 | (57.8-76.8] | 66.1 | (56.2-74.7] | 50.9 | (40.8-60.9] |
| Takeo | 55.5 | (47.8-62.9] | 53.2 | (41.5-64.5] | 54.6 | (45.4-63.4] |
| Odar Meanchey | 73.4 | (57.5-84.9] | 60.0 | (51.9-67.7] | 63.1 | (55.4-70.1] |
| Battambong/Pailin | 54.6 | (47.2-61.8] | 55.1 | (46.7-63.3] | 50.1 | (38.6-61.6] |
| Kampot/Kep | 48.3 | (41.3-55.3] | 51.7 | (41.5-61.8] | 59.7 | (50.5-68.3] |
| Sihanoukville/Koh Kong | 74.7 | (66.7-81.3] | 58.8 | (49.7-67.4] | 57 | (49.7-63.9] |
| Preah Vihear/Stung Treng | 67.5 | (60.3-73.9] | 54.7 | (48.2-61.1] | 68.6 | (61.1-75.2] |
| Mondulkiri/Rattanakiri | 62.6 | (56.3-68.5] | 53.1 | (47.1-59.1] | 57.1 | (48.4-65.5] |
| Overall | 62.2 | (59.8-64.6] | 55.8 | (53.5-58.1] | 56.6 | (54.4-58.7] |
